# Imaging Clusters of Pediatric Low-Grade Glioma are Associated with Distinct Molecular Characteristics

**DOI:** 10.1101/2024.12.16.24319099

**Authors:** Anahita Fathi Kazerooni, Adam Kraya, Komal S. Rathi, Meen Chul Kim, Varun Kesherwani, Ryan Corbett, Arastoo Vossough, Nastaran Khalili, Deep Gandhi, Neda Khalili, Ariana M. Familiar, Run Jin, Xiaoyan Huang, Yuankun Zhu, Alex Sickler, Matthew R. Lueder, Saksham Phul, Phillip B. Storm, Jeffrey B. Ware, Jessica B. Foster, Sabine Mueller, Jo Lynne Rokita, Michael J. Fisher, Adam C. Resnick, Ali Nabavizadeh

**Affiliations:** Center for Data-Driven Discovery in Biomedicine (D3b), The Children’s Hospital of Philadelphia, Philadelphia, PA, USA; AI2D Center for AI and Data Science for Integrated Diagnostics, University of Pennsylvania, Philadelphia, PA, USA; Department of Neurosurgery, Perelman School of Medicine, University of Pennsylvania, Philadelphia, PA, USA; Department of Neurosurgery, The Children’s Hospital of Philadelphia, Philadelphia, PA, USA; Division of Oncology, The Children’s Hospital of Philadelphia, Philadelphia, PA, USA; Division of Radiology, Children’s Hospital of Philadelphia, Philadelphia, PA, USA; Department of Radiology, Perelman School of Medicine, University of Pennsylvania, Philadelphia, PA, USA; Department of Pathology and Laboratory Medicine, The Children’s Hospital of Philadelphia, Philadelphia, PA, USA; Department of Pediatrics, University of Pennsylvania Perelman School of Medicine, Philadelphia, PA, USA; Department of Neurology and Pediatrics, University of California San Francisco, San Francisco, CA, USA; Center for Cancer and Immunology Research, Children’s National Hospital, Washington, DC, USA; Department of Pediatrics, The George Washington University, Washington, DC, USA

**Keywords:** Pediatric low-grade glioma, MRI, Transcriptomic, Genomic, Radiogenomic

## Abstract

**Background:** Cancers show heterogeneity at various levels, from genome to radiological imaging. This study aimed to explore the interplay between genomic, transcriptomic, and radiophenotypic data in pediatric low-grade glioma (pLGG), the most common group of brain tumors in children.

**Methods:** We analyzed data from 201 pLGG patients in the Children’s Brain Tumor Network (CBTN), using principal component analysis and K-Means clustering on 881 radiomic features, along with clinical variables (age, sex, tumor location), to identify imaging clusters and examine their association with 2021 WHO pLGG classifications. To determine the transcriptome pathways linked to imaging clusters, we employed a supervised machine learning model with elastic net logistic regression based on the pathways identified through gene set enrichment and gene co-expression network analyses.

**Results:** Three imaging clusters with distinct radiomic characteristics were identified. *BRAF V600E* mutations were primarily found in imaging cluster 3, while *KIAA1549::BRAF* fusion occurred in subtype 1. The model’s predictive accuracy (AUC) was 0.77 for subtype 1, 0.78 for subtype 2, and 0.70 for subtype 3. Each imaging cluster exhibited unique molecular mechanisms: subtype 1 was linked to oxidative phosphorylation, *PDGFRB*, and interleukin signaling, whereas subtype 3 was associated with histone acetylation and DNA methylation pathways, related to *BRAF V600E* pLGGs.

**Conclusions:** Our radiogenomics study indicates that the intrinsic molecular characteristics of tumors correlate with distinct imaging subgroups in pLGG, paving the way for future multi-modal investigations that may enhance understanding of disease progression and targetability.

## INTRODUCTION

Cancer is a complex biological system influenced by a tumor’s evolutionary environmental forces. Tumor phenotypes are shaped by a combination of interconnected molecular events across the genome, transcriptome, epigenome, and proteome among other cellular contexts, which collectively converge to dysregulate cellular biological functions in ways that promote oncogenesis and cancer progression ^1^. Exploring the relationships between radiophenotypes—tumor traits beyond conventional analysis quantified from radiological imaging that reflect characteristics at genomic and transcriptomic levels—can enhance our understanding of tumor dynamics and the variability in therapeutic responses across similar histologies ^2^.

Pediatric low-grade gliomas (pLGGs), the most common brain tumors in children, account for one-third of all pediatric brain cancers ^3, 4^. While complete resection improves survival, deeply-seated or infiltrative tumors often necessitate chemotherapy post-partial resection, leading to a 10-year event-free survival (EFS) of around 50% ^5, 6^. Treatments can also impact cognitive and neurological outcomes in longer term, affecting quality of life and survivorship ^7^.

PLGGs encompass a wide array of molecular subtypes, each associated with different prognoses and responses to treatment, underscoring the need for treatments targeted to specific subtypes. The introduction of targeted treatments, including *RAF* or *MEK* inhibitors, has expanded treatment options for patients with pLGG, paving the way for improved patient outcomes ^8, 9^. However, understanding the biological and molecular basis of pLGG tumors beyond single gene mutation is critical for the success of these targeted therapies and may be helpful to address current challenges of tumor recurrence, rebound, and developing resistance in the era of molecularly targeted agents ^8, 10^.

Radiomics – a high throughput approach for extracting quantitative features from radiological images – offers the potential to provide non-invasive biomarkers for characterizing the molecular underpinnings of tumors ^11, 12^. However, most existing studies on the radiogenomic analysis of pLGGs focus on predicting individual gene alterations, such as *KIAA1549::BRAF* fusion or *BRAF V600E* mutation ^13–16^. As WHO CNS 5 classification recommends the inclusion of driver molecular alterations in standard diagnosis, predicting a single mutation or fusion using imaging alone may not be useful or sufficient to fully understand the tumor biology. Moreover, focusing on a single pathway or gene mutation limits our understanding of the complex interactions between different pathways that drive patient-specific responses to targeted treatments.

We have formerly explored the association of imaging clusters, referred to as imaging subtypes, derived through an unsupervised machine learning approach based on radiomic features, with key genomic characteristics in pLGG ^17^. Unsupervised clustering based on imaging characteristics aims to uncover distinct and relatively homogeneous subgroups or subtypes within a patient population. This approach facilitates the identification of patterns and associations that may not be apparent through traditional analysis methods ^18^. Our hypothesis is that radiological patterns – quantified using radiomic features - integrated with machine learning, can uncover molecular aspects of the tumor and potential treatment targets, beyond molecular subtypes. In this radiogenomic study, we intend to elucidate how information from genomic and transcriptomic scales translates into radiophenotypic characteristics.

## METHODS

The overall workflow of the analysis is illustrated in the graphical representation shown in Figure 1.

**Figure 1.**
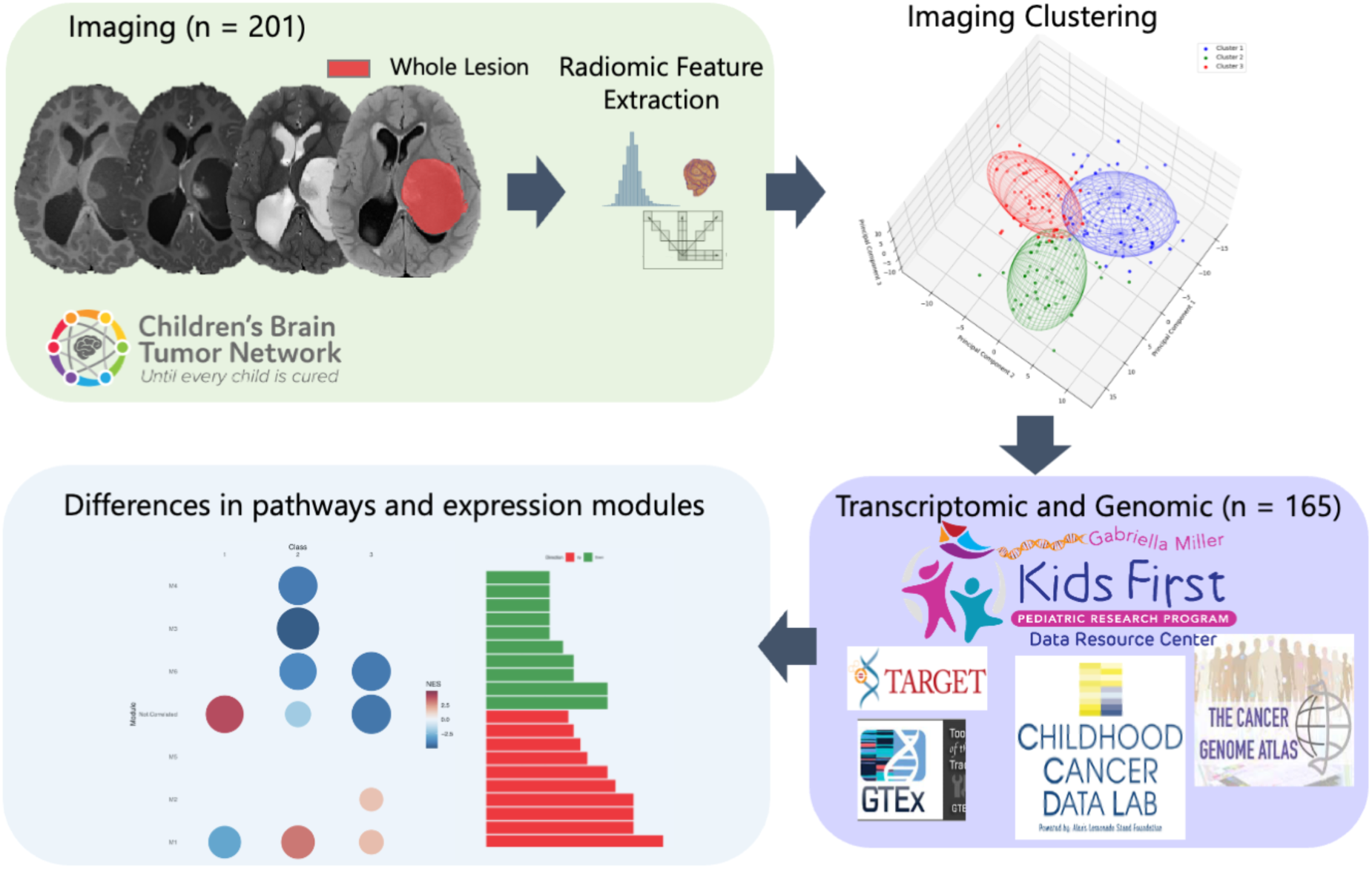
Graphical representation of the analysis steps.

### Overview of the Data

In this study, we analyzed retrospectively-collected data from the Children’s Brain Tumor Network (CBTN) ^19^ repository (cbtn.org), which includes specimens and longitudinal clinical and imaging data, facilitating the sharing of de-identified samples for research. The study complied with HIPAA guidelines and received IRB approval from the Children’s Hospital of Philadelphia (CHOP) through the CBTN protocol, with informed consent obtained for patient enrollment.

We focused on patients histopathologically diagnosed with *de novo* pLGG from 2006 to 2018. Standard multiparametric MRI (mpMRI) sequences, including pre- and post-Gadolinium T1-weighted (T1w, T1w-Gd), T2-weighted (T2w), and T2 fluid-attenuated inversion recovery (T2-FLAIR), were collected from 258 pLGG subjects. Exclusion criteria included patients older than 18 years at the time of imaging, imaging conducted post-surgery or treatment, tumors outside the brain, leptomeningeal dissemination, missing MRI sequences, or low-quality imaging. After exclusions, 201 subjects with complete mpMRI sequences were included for analysis. Molecular data from 165 intersecting subjects were compiled from CBTN records and the Open Pediatric Cancer (OpenPedCan) project repository (version 15 data release; DOI: <u>10.5281/zenodo.6473912</u>) ^20^. Patient characteristics are summarized in Supplementary Table 1.

### Imaging Data Analysis

Details on image preprocessing are provided in Supplementary Material SI1. Images underwent skull stripping ^21^ and intensity normalization to a scale of [0, 255] after removing outliers to facilitate radiomic feature extraction. We generated Whole Tumor (WT) masks by combining all four tumor components generated using our in-house automatic pediatric brain tumor segmentation tool (https://github.com/d3b-center/peds-brain-auto-seg-public) ^21, 22^, followed by manual revisions when necessary. An extensive array of radiomic features (n = 881), including metrics of shape, volume, intensity, and texture were extracted. The texture analyses included techniques such as gray-level co-occurrence matrix (GLCM), gray-level run-length matrix (GLRLM), gray-level size zone matrix (GLSZM), neighborhood gray tone difference matrix (NGTDM), local binary pattern (LBP), and Collage ^23^ features, using the WT masks on the multiparametric MRI images.

We randomly split the dataset into a discovery group (80% of the cohort; n = 160) for model training, and a replication group (20% of the cohort; n = 41) for testing, and conducted a pairwise Pearson’s correlation analysis on the radiomic features within the discovery group, eliminating one feature from each pair that had a correlation coefficient (r) exceeding 0.90, reducing the number of features to 438. These features were normalized via z-scoring. Tumor location, a categorical variable, was normalized through count encoding, and age at diagnosis was standardized using z-scoring based on the mean and standard deviation of continuous features from the discovery group, which was then applied to normalize the features in the replication group. We produced histograms that display the distribution of signal intensities within specific sequences (x-axis) against their frequency (y-axis) across the entire dataset, providing insights into the characteristic distribution of these features among different pLGG imaging clusters.

### Imaging-Based pLGG Clusters

We applied a multi-step clustering approach to a feature set comprising 441 variables, including 438 radiomic features, along with age, sex, and tumor location. To reduce the dimensionality of the feature space, we utilized principal component analysis (PCA) and retained 78 principal components (PCs), capturing over 95% of the total variance and preserving the essential information required for accurate modeling and analysis. To determine the optimal number of imaging-based pLGG clusters, we used the “elbow method”, which analyzes the “within-cluster sum of squares” across the number of clusters ranging from 1 to 160, identifying the point of diminishing returns. To ensure robust cluster determination, we supplemented the elbow method with the “silhouette coefficient”, calculated for cluster counts from 2 to 15. This metric, ranging from -1 to 1, validated the optimal cluster number by maximizing inter-cluster separation and intra-cluster cohesion.

After identifying the optimal number of clusters, we used the “Greedy K-means++” algorithm to cluster the PCs from the discovery cohort. This method, which is less sensitive to initial centroid selection, improves clustering reliability and stability. To mitigate any potential biases from random initialization and ensure reproducibility, the model was trained 1,000 times in a single run. The finalized model was then used to predict cluster membership for the replication cohort (*n* = 41), to evaluate generalizability.

### Differential Gene and Pathway Expression Analyses

We analyzed clinical and RNA-Seq data from the CBTN ^19^ repository using the STAR-RSEM RNASeq pipeline of the OpenPBTA ^20^. Details on gene set enrichment analysis (GSEA) can be found in Supplementary Material SI2. To identify pathway expressions that are most predictive of imaging cluster assignment, using the glmnet R package, we trained an ElasticNet logistic regression classifier using a one-versus-rest strategy for each imaging cluster, leveraging 70% of the low-grade glioma cohort in this study to train – validate the model and leaving 30% as an independent test set, and used 10-fold cross validation to tune the alpha and lambda model parameters. We selected the top 100 most variable differentially expressed/co-expressed pathways along with molecular subtype as described by Ryall et al ^6, 20^, the anatomical region of the tumor, age at diagnosis, reported sex, and race as predictors. We evaluated the area under the receiver operating curve (AUC-ROC) of the training – validation and independent testing sets to evaluate the degree of overfitting for each model and retained pathways with non-zero generalized linear model (GLM) coefficients for further evaluation.

### Association Analysis between Imaging and pLGG WHO 2021 Classification

PLGG tumor entities described by the 2021 World Health Organization (WHO) classification of central nervous system (CNS) tumors ^4^ were compiled based on molecular and pathological information from the OpenPedCan v15 data repository (described before), which annotates pLGG with well-defined genomic lesions, including single nucleotide (SNVs), insertion-deletion (indel), and structural variants (SVs) occurring in MAPK pathway-related and non-MAPK pathway-related genes. The association between imaging and WHO 2021 classifications was evaluated using a likelihood ratio test (LRT) between a saturated Poisson GLM and a nested Poisson model excluding the interaction term. A correspondence analysis was then performed on the Pearson residuals derived from the Poisson saturated model to visualize the associations between imaging and pLGG WHO CNS tumor classifications.

### Analysis of *KIAA1549::BRAF* Fusion Breakpoints

All high-confidence, in-frame *KIAA1549*::*BRAF* STAR-fusion and/or Arriba fusion calls in pLGG tumors were annotated with exon number in canonical transcripts (NM_001164665 for *KIAA1549* and NM_004333 for *BRAF*) using *biomaRt* and *GenomicRanges* R packages ^24–26^. Common breakpoints included those involving exons 15:09 (exon 15 in *KIAA1549* and exon 9 in *BRAF*), 16:09, 16:11, and 18:10 ^6, 27^. All other breakpoint combinations were classified as rare/novel.

### Data and Code Availability

The processed data and the codes utilized for the analysis of genomic, transcriptomic, and radiomic data in conjunction with patient clinical characteristics is accessible at https://github.com/d3b-center/pLGG_imaging_clustering_genomic. All image processing tools employed in this study are publicly available and free to use, including CaPTk (https://www.cbica.upenn.edu/captk) and the in-house automated tumor segmentation model (https://github.com/d3b-center/peds-brain-auto-seg-public).

## RESULTS

Our clustering model stratified patients into three imaging clusters (Figure 2A, Supplementary Figure 1) and was applied to the replication set. Histograms (Supplementary Figure 2), presenting variations in imaging characteristics across the three imaging clusters, showed cluster 2 had moderate contrast enhancement based on the location of the peak of histogram falling around intermediate intensity values. This cluster also shows higher peak of T2-FLAIR intensity values within peritumoral edema in the higher range of intensity values. Additionally, reduced T2 intensity values were observed in the non-enhancing tumor regions for imaging cluster 2 compared to the other clusters, which might suggest a denser tumor cell population.

**Figure 2.**
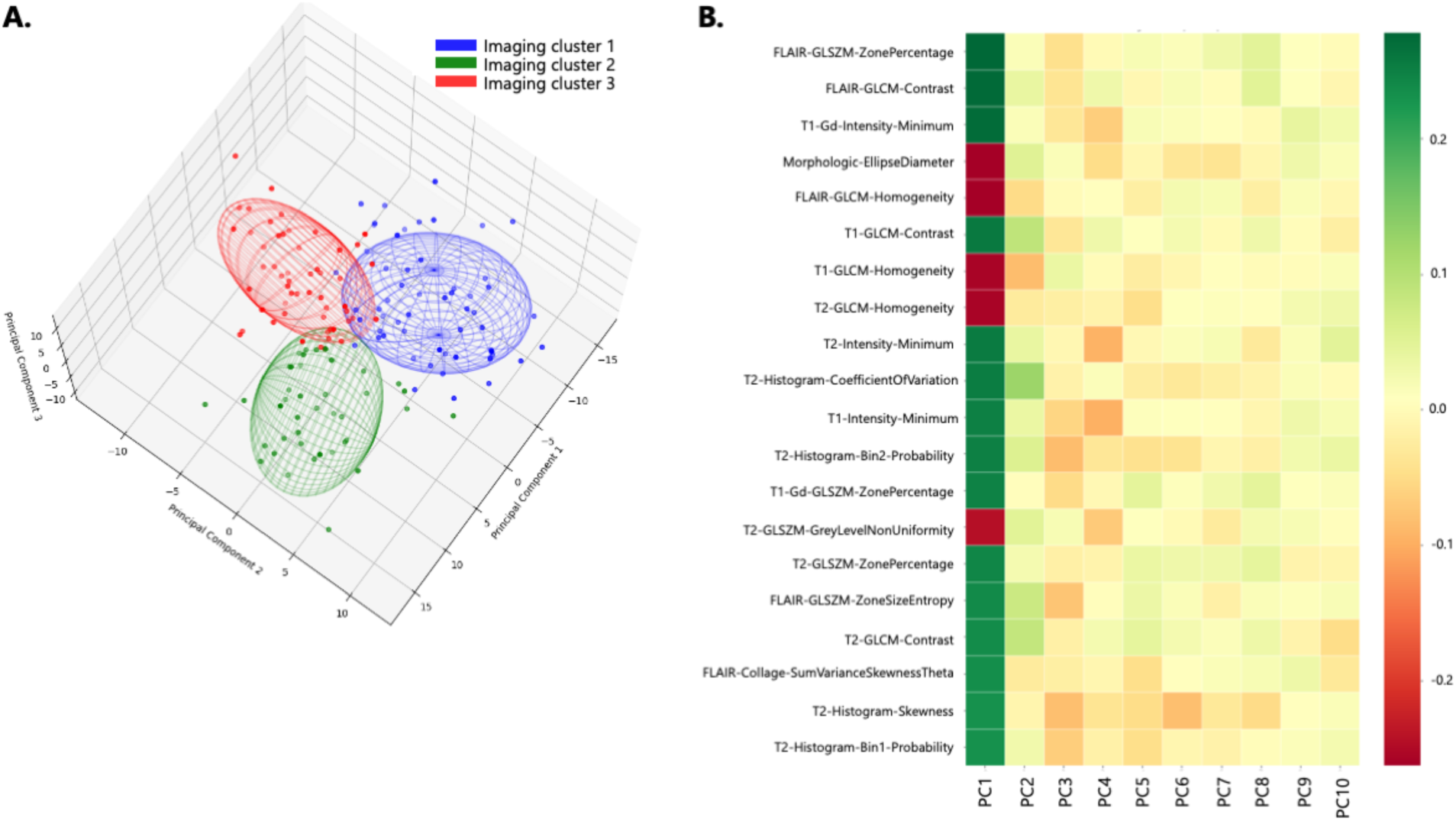
(A) Three-dimensional visualization of imaging clusters in the discovery set; (B) Heatmap plot of the radiomic features contributing to the top 10 principal components (PCs).

Top radiomic features influencing PCs (Figure 2B) mainly included the texture features that reflect tumor heterogeneity, such as GLCM contrast and homogeneity features and the coefficient of variation and skewness from the first-order histogram feature family. Additionally, a morphologic feature was also identified as a key contributor to the composition of the PCs.

We examined the relationship between imaging clusters and 2021 pLGG WHO CNS tumor classifications ^4^. Correspondence analysis of Pearson residuals from a Poisson GLM revealed pilocytic astrocytomas were over-represented in cluster 1 (Figure 3A), while cluster 2 was enriched with diffuse low-grade glioma, MAPK-altered tumors (IRR = 4.67, p<0.001), and gangliogliomas (IRR = 11.2, p = 0.022), but under-represented pilocytic astrocytomas (IRR = 0.6, p = 0.034). Cluster 3 also under-represented pilocytic astrocytomas (IRR = 0.5, p<0.001), and included MAPK-altered diffuse low-grade glioma tumors (IRR = 2.85, p<0.001) and pediatric-type diffuse low-grade gliomas, NOS (Figure 3A). The alluvial plot in Figure 3B shows the association between imaging clusters and tumor location. In tandem with Figure 3A, cluster 1 predominantly included cerebellar pilocytic astrocytomas with *KIAA1549::BRAF* fusions, as observed in other studies ^9, 28^, while cluster 3 was linked to diffuse low-grade gliomas with *BRAF V600E* mutations across various brain regions.

**Figure 3.**
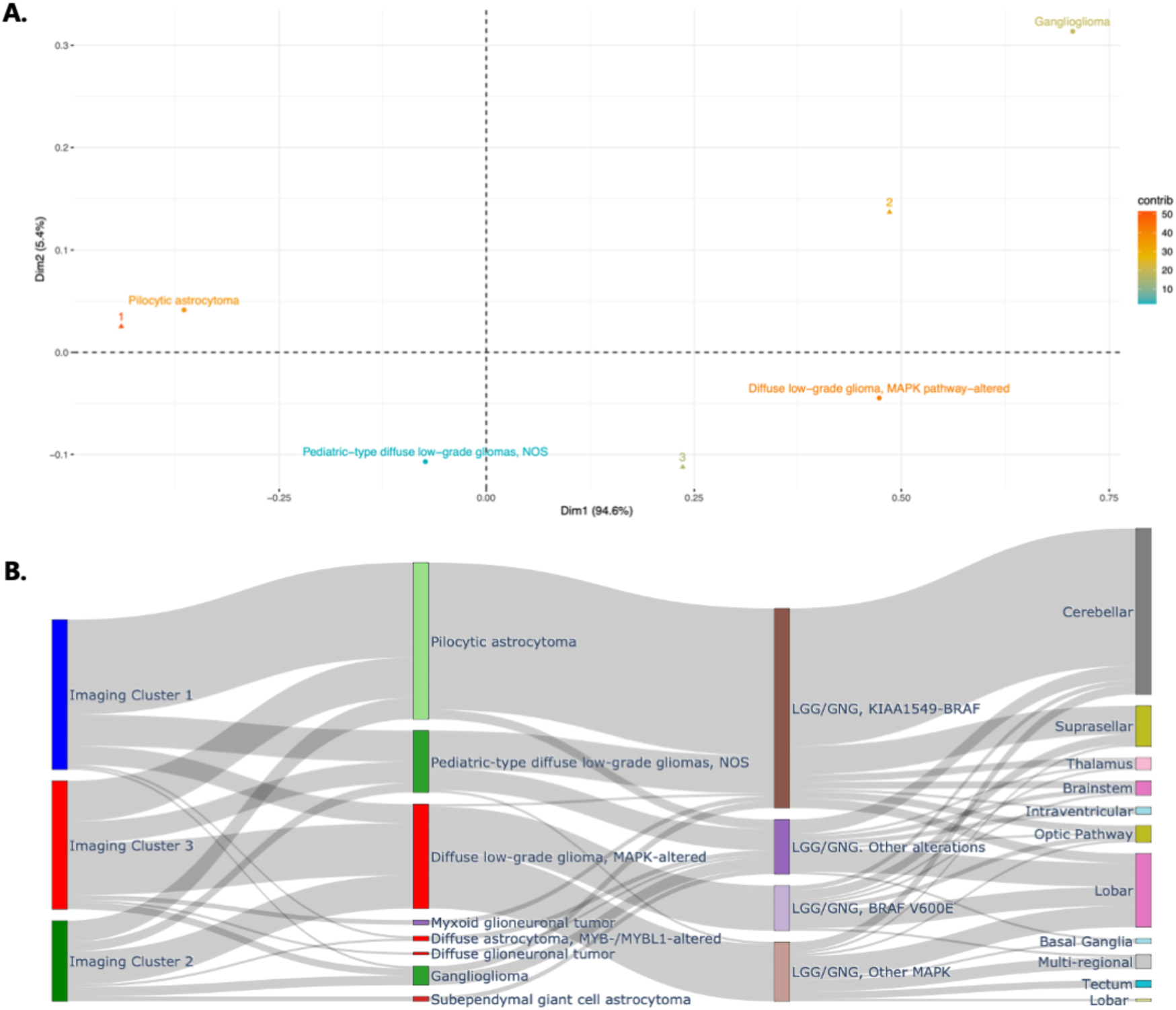
(A) correspondence analysis plot of Pearson residuals depicting the association between imaging clusters and pLGG 2021 WHO CNS Tumor Classifications. (B) Alluvial plot indicating associations of imaging clusters with pLGG 2021 WHO CNS Tumor Classifications, pLGG molecular subtypes by Ryall et. al, and tumor locations.

Transcriptomic analysis of imaging clusters using GSEA (Figures 4A-C) revealed distinct pathway expressions. Cluster 1 showed lower expression of fatty acid metabolism pathway, critical for glioma growth ^29, 30^, and rhodopsin-like G-Protein coupled receptor (*GPCR*) signaling ^31^, and higher expression of PDGFRB-related and oxidative phosphorylation pathways compared to cluster 2 ^5, 32^. Compared to cluster 3, cluster 1 also showed reduced expression of rhodopsin-like receptors and *FGFR1* signaling, frequently mutated in midline low grade gliomas ^33, 34^, fewer *FGFR1* mutations and a cerebellar location. Pathways with higher expression in cluster 1 versus cluster 3 included oxidative phosphorylation and immune regulatory pathways (complement cascade, MHC class I antigen presentation). Comparing clusters 2 and 3, higher expression in adhesion-related and extracellular matrix remodeling pathways were found, suggesting differences in migratory potential.

**Figure 4.**
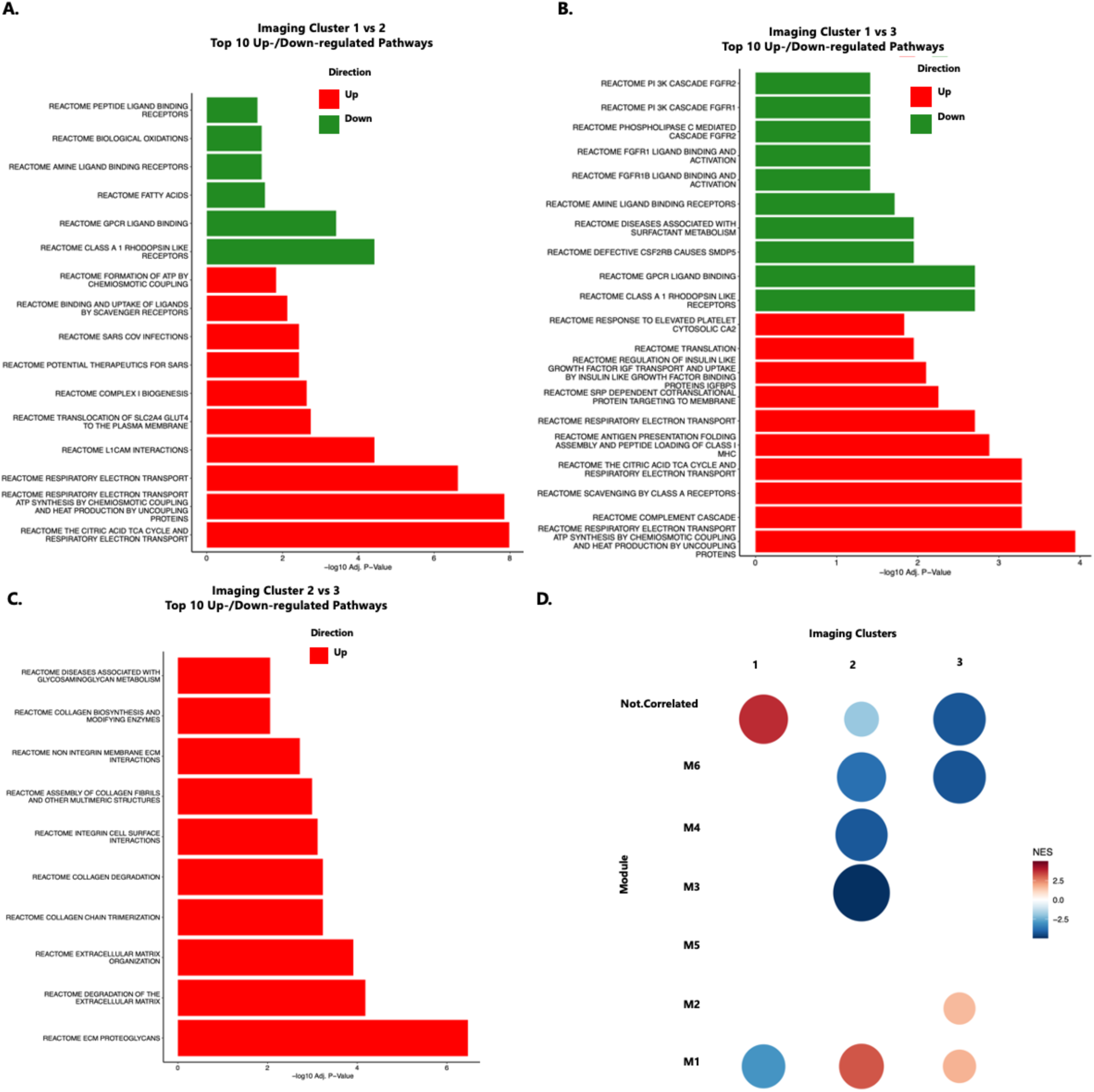
Molecular pathways associated with imaging clusters: (A-C) Top GSEA pathways differentially up-regulated or down-regulated between pairs of imaging clusters; (D) matrix of Pearson’s residuals for co-expression modules (defined by CEMITool) across the three imaging clusters.

Gene co-expression analysis identified seven differentially expressed modules across imaging clusters (CEMITool, see Supplementary Material SI2; Figure 4D). Consistent with GSEA, lower expression of rhodopsin-like GPCRs and potassium channel signaling was observed in cluster 1 relative to clusters 2 and 3 for module M1, along with GABA signaling and synaptic neurotransmission. Module M2 showed higher activity in cluster 3 for histone acetylation and DNA methylation pathways. Module M3 showed decreased activity in cluster 2 for potassium signaling, largely non-overlapping with those of Module M1, and glucose homeostatic pathways involving glucagon and secretin hormones. Module M4 displayed lower interleukin (ILs 4, 6, 10, 13) and PD-1 signaling levels in cluster 2. Module M6 demonstrated lower extracellular matrix remodeling and integrin-related pathway expression in clusters 2 and 3, with cluster 3 exhibiting the lowest expression, consistent with GSEA findings. Non-correlated genes were enriched in tumor-promoting interleukin pathways (ILs 4, 10, and 13 ^35–38^), particularly in cluster 1. Module M5 showed no significant differences or biological enrichment across clusters.

Leveraging a machine learning-based approach to compare differentially expressed/co-expressed signaling pathways, molecular subtypes, and clinical variables across imaging clusters resulted in an average AUC-ROC of 0.83 | 0.77 (training balanced accuracy: 0.79) when predicting cluster 1, 0.94 | 0.78 when predicting cluster 2 (training balanced accuracy: 0.84), and 0.84 | 0.70 for training-validation | testing sets respectively when predicting cluster 3 (training balanced accuracy: 0.8; ROC curves are depicted in Supplementary Figure 3). Figure 5A-C illustrates GLM coefficients for each classifier. Tumor anatomical location significantly influenced cluster 1 classification (Figure 5A). Collagen degradation pathways and protein-protein interactions at neuronal synapses were the most positively and negatively influential pathways associated with cluster 1, respectively. For cluster 2 (Figure 5B, in line with Figure 3A-B), diverse molecular subtypes were identified, including deleterious alterations in *BRAF* that were negatively predictive. In contrast to cluster 1, protein-protein interactions at neuronal synapses were positively associated with cluster 2 (Figure 5B); transcriptional regulation by the hematopoiesis regulator *RUNX1* and non-integrin ECM interactions were positively associated with cluster 2. In addition to BRAF-associated subtypes, we identified *GPCR* signaling positively associated with cluster 3 and notably non-homologous end joining-based DNA repair negatively associated (Figure 5C).

**Figure 5.**
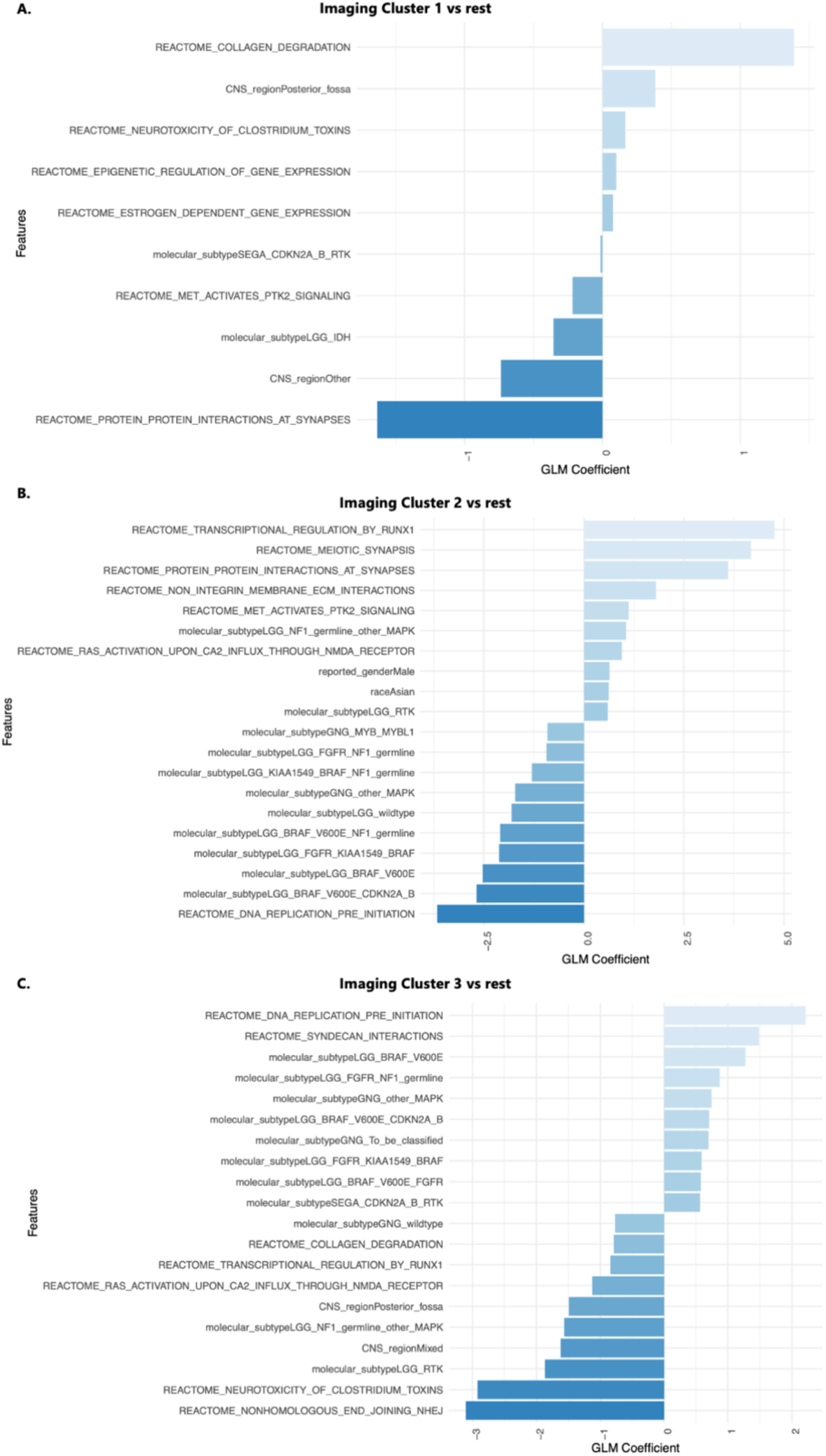
(A-C) Plots of generalized linear model (GLM) coefficients indicating the top pathways selected in the elastic net classification approach for prediction of each imaging cluster vs the rest of the tumor cohort.

Kaplan-Meier survival analysis and log-rank tests did not show significant survival differences across clusters (p > 0.05). However, Cox regression analysis (Figure 6A), adjusting for extent of tumor resection, molecular subgroups, and imaging clusters as covariates, revealed a significantly better prognosis for cluster 3 compared to cluster 1 (used as the reference group). Interestingly, an interaction analysis between tumors exhibiting *KIAA1549::BRAF* fusion and imaging cluster 3 suggested a less favorable prognosis for this subtype of tumors in cluster 3 (HR = 47, p = 0.012) (Figure 6A). We observed *BRAF*-fusion status-dependent differences in patient survival in cluster 3 (Supplementary Figures 4-7). Among patients with non-BRAF fusion tumors, those in cluster 3 exhibited significantly better EFS relative to cluster 1 in a Cox regression model that included covariates for extent of tumor resection (HR = 0.22, p = 0.002; Supplementary Figure 5). However, there was no survival benefit for patients in cluster 3 relative to cluster 1 among patients with *BRAF*-fusion tumors (HR = 1.3, p = 0.573; Supplementary Figure 7).

**Figure 6.**
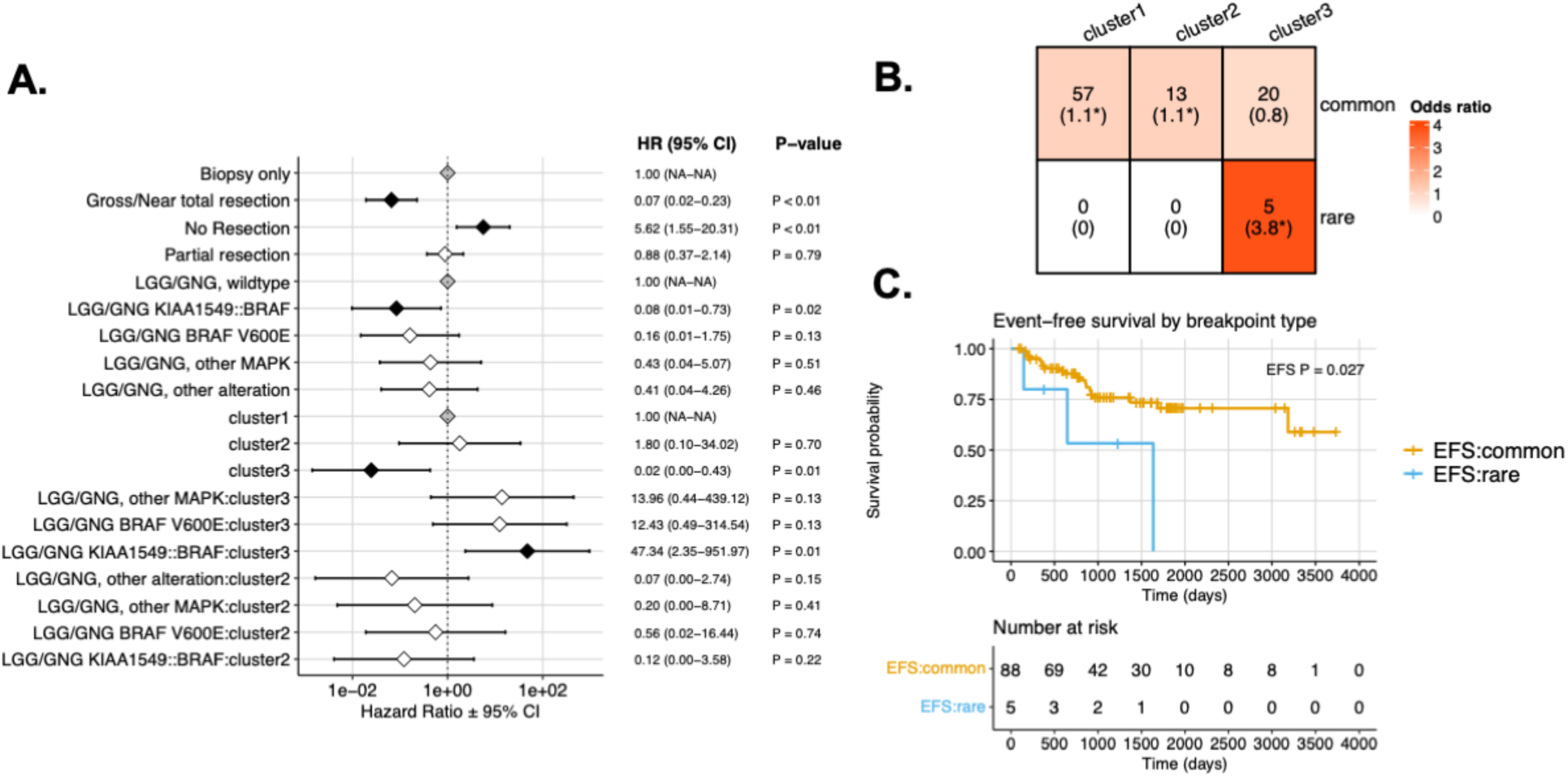
Event-free survival (EFS) by imaging cluster. (A) Cox regression model forest plot of EFS, including covariates for extent of tumor resection, molecular subtype, imaging cluster, and subtype-imaging cluster interaction terms. Gray points indicate reference levels (Biopsy only, LGG/GNG wildtype, and imaging cluster 1), and black points indicate terms with statistically significant hazard ratios relative to reference levels. (B) Distribution of KIAA1549::BRAF fusion tumor breakpoint types and corresponding enrichment odds ratios in parentheses. *p<0.05. (C) Kaplan-Meier plot of EFS among patients with KIAA1549::BRAF fusion tumors by KIAA1549::BRAF breakpoint type.

We identified *KIAA1549::BRAF* fusion breakpoint exons to determine their distribution in each imaging cluster. Following previous work ^27^, we defined common exon breakpoints as those involving exons 15:09 (exon 15 in KIAA1549 and exon 9 in BRAF), 16:09, 16:11, and 18:10, while all other breakpoint combinations were classified as rare. Breakpoint types (common vs. rare) were significantly associated with imaging cluster (Fisher exact p = 2.0e-03), with rare *KIAA1549::BRAF* fusion breakpoint tumors found solely in imaging cluster 3 (Figure 6B, Supplementary Figure 8). Patients with *KIAA1549::BRAF* rare breakpoint tumors have previously been shown to exhibit worse EFS relative to those with common breakpoints. We recapitulated this finding in our cohort in a log-rank test (p = 0.03, Figure 6C). However, there was no significant difference in EFS between patients with rare vs. common breakpoints in a Cox regression model that included covariates for extent of tumor resection and imaging cluster (HR = 1.7, p = 0.48; Supplementary Figure 9).

## DISCUSSION

In this manuscript, we presented an imaging-based clustering method for pLGGs and identified three subgroups with different molecular characteristics, beyond what is captured via characterization of classic molecular subtypes. Our analysis revealed distinct transcriptomic profiles across different imaging clusters, providing deeper insights into the molecular underpinnings of glioma behavior and potential future therapeutic targets. Imaging cluster 1 showed unique mechanisms that may drive tumor growth, including up-regulation of oxidative phosphorylation, *PDGFRB*, and tumor-promoting interleukin 4, 10, and 13 signaling ^35, 39, 40^. Notably, microglia have been found to induce *PDGFRB* expression in low-grade glioma to enhance migratory capacity ^5, 32^. *KIAA1549::BRAF* pilocytic astrocytomas enriched in cluster 1 have been found to aberrantly activate oxidative phosphorylation in part through deleterious mitochondrial DNA mutations ^41, 42^. Elevated IL10 levels, typically found in invasive higher grade gliomas that suppress anti-tumor immune activity, highlight potential for targeted therapies with direct inhibitors or neutralizing antibodies ^35^. Cluster 2 showed higher class A rhodopsin-like receptor signaling and fatty acid oxidation, features associated with metabolic plasticity and glioma stem cell maintenance, common in higher-grade gliomas ^31^. These findings highlight the therapeutic potential of inhibiting glycolytic and fatty acid oxidation pathways ^29, 30, 43^.

Differential expression in adhesion and extracellular matrix remodeling pathways between clusters 2 and 3 suggests variations in microenvironmental remodeling, potentially influencing tumor spread, invasiveness, immune escape, and response to immune checkpoint blockade ^44^. A particularly intriguing finding was the up-regulation of pathways related to histone acetylation and DNA methylation in cluster 3, which may be tied to the enrichment of *BRAF V600E* low grade gliomas ^45^. Epigenetic reprogramming, observed in *BRAF* V600E-driven melanomas and colorectal cancers, enables resistance to *BRAF* inhibitors overcomes *BRAF* dependency, counters oncogene-induced senescence, and overcomes *BRAF* dependency, highlighting the need for combination therapies targeting BET proteins or histone deacetylases alongside the oncogenic *BRAF* mutation ^45–47^. Additionally, compared to cluster 1, cluster 3 exhibited elevated *FGFR1* signaling, underscoring its distinct molecular features.

In our analysis of *KIAA1549::BRAF* fusion breakpoints across the three imaging clusters, the novel/rare breakpoints (*KIAA1549::BRAF* 15:11) in pLGGs were all classified into cluster 3, indicating similar imaging characteristics among these patients. Consistent with previous studies ^6, 27^, tumors with rare/novel *KIAA1549::BRAF* fusion breakpoints, which are highly enriched in recurrent/progressive pLGGs, exhibit a poorer prognosis. However, these patients with poorer survival outcomes were among those in imaging cluster 3, which is associated with a more favorable prognosis. Furthermore, in cluster 3, we observed a higher hazard ratio for *KIAA1549::BRAF* tumors compared to tumors with *BRAF V600E* mutation. Although the *BRAF V600E* mutation did not yield a significant hazard in Cox regression analysis (p = 0.13), this finding contrasts with the established poorer prognosis of pLGGs with the *BRAF V600E* mutation compared to those with *KIAA1549::BRAF* fusion ^48^. This discrepancy may be due to the inclusion of rare or novel *KIAA1549::BRAF* fusion breakpoints in cluster 3, and the fact that 8 out of 12 tumors with the *BRAF V600E* mutation in this group had not progressed during the time period of this study. These findings highlight the need for a larger study to identify more granular clusters with better differentiation across higher and lower risk tumors.

The current literature on radiomic analysis pLGG is limited, primarily focusing on predicting single genetic mutations or alterations, such as the *BRAF V600E* mutation and *KIAA1549::BRAF* fusion, using MRI ^13–16^. However, as the 2021 WHO CNS 5th Edition recommends incorporating driver molecular alterations into diagnostics ^4^, predicting a single mutation or fusion using imaging alone may have limited clinical utility. Furthermore, these studies have not explored associations between imaging phenotypes and underlying genomic or transcriptomic pathways.

Our study offers the first comprehensive radiogenomic analysis investigating genomic alterations and pathways underlying specific imaging characteristics in a group of patients. While this approach is unexplored in pediatric neuro-oncology, identifying imaging-based subgroups through unsupervised clustering and linking them to clinical symptoms, treatment responsiveness, and genomic factors has been applied in neurodegenerative diseases and adult glioblastoma ^18^.

For instance, one study ^49^ identified three glioblastoma (GBM) subtypes through unsupervised clustering, revealing differences in outcomes and molecular features such as *IDH1* status, *MGMT* promoter methylation, *EGFRvIII* expression, and transcriptomic subtypes. Another study ^50^ utilized a radiomic-genomic joint learning approach to define three GBM imaging subtypes with distinct survival probabilities and gene mutation patterns.

Our study faced challenges due to the limited availability of imaging and genomic data (WGS and RNA-Seq), compounded by the rarity of pediatric brain tumors and a small cohort size, particularly for non-pilocytic astrocytomas in specific brain sites. Survival analysis relied on subjective progression assessments from clinical notes and MRI reviews. Supervised machine learning models (ElasticNet logistic regression) showed higher performance on the training set than the test set, likely due to the small sample size and variability in tumor location and molecular subtypes between the training and test sets. The expression data, representing a localized tumor portion may not fully reflect the tumor’s broader molecular heterogeneity, limiting its correlation with radiomic features.

Additionally, other critical data layers, such as methylation profiling, metabolomics, proteomics, and pathomics, were unavailable for most patients. Future efforts should focus on larger cohorts and integrating additional data types to capture the complex interplay between these layers, enhance tumor subtype differentiation, and identify potential therapeutic targets for personalized treatment.

In conclusion, our work showed that unsupervised analysis of treatment-naïve imaging and clinical tumor data available at diagnosis can identify potential candidates for targeted drug selection, extending beyond the WHO-recognized molecular subtypes. This approach enables customization of treatments to match the unique characteristics and needs of each patient, thereby improving the quality and effectiveness of care for patients with pLGGs. Integration of multi-modal data aligns with the advancements in personalized medicine, providing a comprehensive view of each patient’s tumor. This supports molecular tumor boards on pLGGs in developing more targeted treatment strategies, moving beyond conventional methods.

