## Supplementary Material for "Imaging Clusters of Pediatric Low-Grade Glioma are Associated with Distinct Molecular Characteristics"

### SI1. Image Preprocessing and Tumor Subregion Segmentation

In this research, we standardized images prior to extracting radiomic features. Initially, MRI scans from each patient were oriented to the left-posterior-superior (LPS) coordinate system. The T1-weighted (T1w), T2-weighted (T2w), and T2-FLAIR, were aligned with the corresponding post-contrast T1w (T1w-Gd) images. These images were then uniformly resampled to a 1 $mm^3$ isotropic resolution using the anatomical SRI24 atlas through the Greedy algorithm, as facilitated by the open-source Cancer Imaging Phenomics Toolkit (CaPTk, version 1.8.1, available at https://www.cbica.upenn.edu/captk) [1,2]. Following this, the images underwent skull stripping using a specialized pediatric brain tissue extraction tool [3]. After removing outlier pixels that exceeded the 99.9th percentile in the image histogram, the skull-stripped images were normalized to an intensity range of [0, 255]. Brain tumor segmentation was conducted with our proprietary pediatric brain tumor segmentation tool [4], complemented by manual adjustments where needed. This tool delineates various tumor subregions, such as the enhancing tumor, non-enhancing tumor, cyst, and edema, which are collectively used to create a complete lesion segmentation, as illustrated in Figure 1.

### SI2. Enrichment Analysis of Molecular Pathways

Differentially expressed genes were used as input for gene set enrichment analysis (GSEA [5]), and differentially regulated pathways were defined as having a Benjamini-Hochberg corrected p-value of less than 0.05. Pathway annotations were derived from Reactome [6] using the Molecular Signatures Database (MSigDb [7]) v2023.1.Hs 74. The ClusterProfiler [8] R package was used to generate visualizations of differentially regulated pathways. To understand differences in gene co-expression networks, we investigated cohort-wide gene-gene correlations subject to a scale-free topology using CEMITool [9], which internally leverages GSEA to determine differences in network module expression as a function of a categorical annotation (in this case, imaging cluster). We also directly investigated differentially co-expressed pathways using Gene Set Net Correlation Analysis (GSNCA [10]), which estimates gene-specific weights based on a gene's cross-correlation with all other genes in a given pathway. We focused on the identification of differentially co-expressed pathways and hub genes, defined as a gene with the greatest weight in a given pathway.

To identify differentially expressed biological pathways across imaging groups, we filtered gene expression data for the 'protein coding' gene type. We then provided expected counts as input to DeSeq2 [11] for differential gene expression analysis, specifying imaging cluster as the factor variable in the model's design. Lists of differentially expressed genes derived from pairwise comparisons across all three imaging clusters were ranked and used as input for pre-ranked GSEA [12] to identify differentially expressed biological pathways. Pathway annotations were derived from Reactome using the Molecular Signatures Database v2023.1.Hs [7].

To characterize and profile gene co-expression sub-networks in low grade gliomas, we used the CEMITool bioconductor package [9] on TPM gene expression data. CEMITool defines co-expressed modules based on the correlation structure and a re-scaled gene-gene adjacency metric to define subnetworks following a scale-free topology. GSEA and over-representation analyses are then used to characterize cluster-specific subnetwork differences.

*Supplementary Table 1. Summary of demographics and clinical characteristics of the pLGG patients included in the imaging cohort (collected through the CBTN data repository)*

| Variable | Value | # of Patients |
|---|---|---|
| *Age Range (months)* | 4.3 – 280.73; Mean, 108.07 | 201 |
| *Sex* | Female | 97 |
| | Male | 104 |
| *Tumor Location* | Basal Ganglia | 2 |
| | Brainstem | 10 |
| | Cerebellar | 86 |
| | Intraventricular | 5 |
| | Lobar (frontal, parietal, temporal, or occipital lobes) | 44 |
| | Multi-regional | 14 |
| | Suprasellar | 34 |
| | Thalamus | 6 |
| *Extent of Tumor Resection* | Gross or near total resection | 109 |
| | Partial resection | 58 |
| | Biopsy | 27 |
| | Not Reported/Unavailable | 7 |
| *Treatment* | Radiotherapy | 10 |
| | Chemotherapy | 46 |
| | None or Not Available | 149 |
| *WHO 2021 Classification* | Diffuse astrocytoma, MYB- or MYBL1-altered | 3 |
| | Diffuse glioneuronal tumor with oligodendroglioma-like features and nuclear clusters | 1 |
| | Diffuse low-grade glioma, MAPK pathway-altered | 128 |
| | Ganglioglioma | 5 |

| | | |
|---|---|---|
| | Glioneuronal and neuronal tumors | 3 |
| | Pediatric-type diffuse low-grade gliomas, NOS | 33 |
| | Pilocytic astrocytoma | 25 |
| | Subependymal giant cell astrocytoma | 3 |
| *Progression-Free Survival Range (months)* | 2.83 – 133.37; Mean, 39.88<br>61 progressed | 201 |

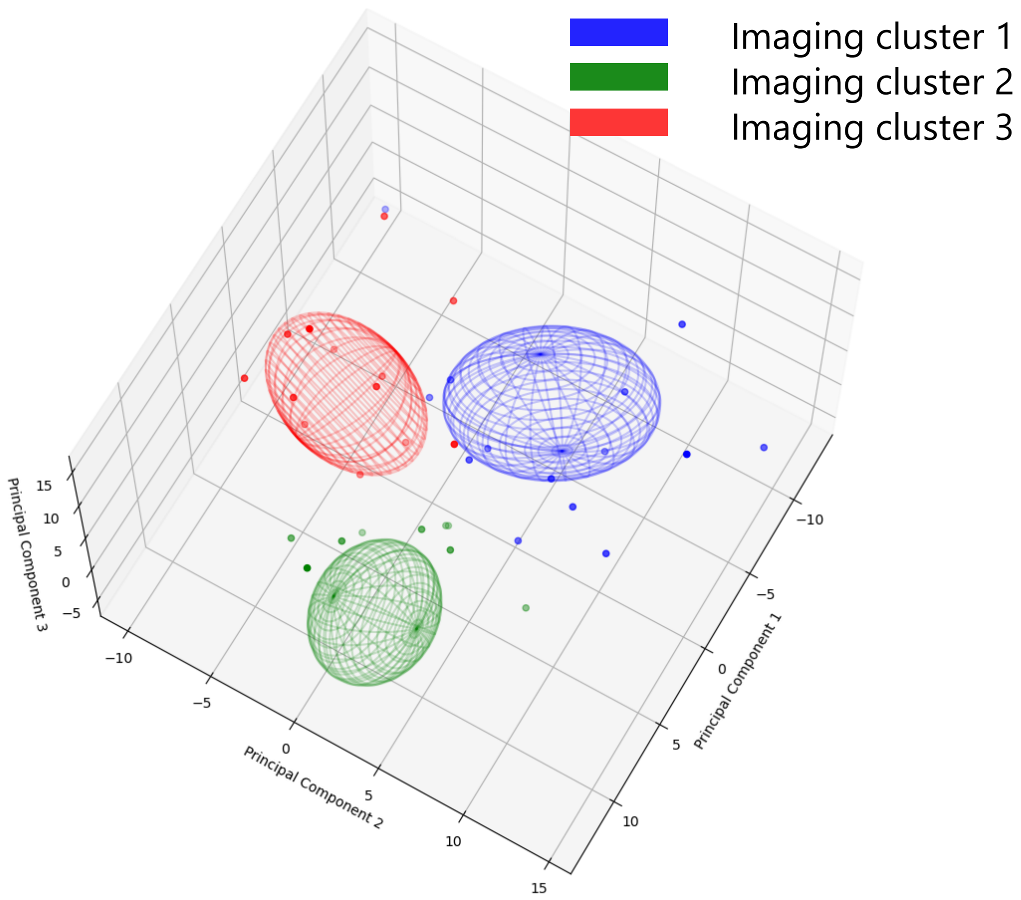


*Supplementary Figure 1. Three-dimensional visualization of imaging clusters in the replication set.*

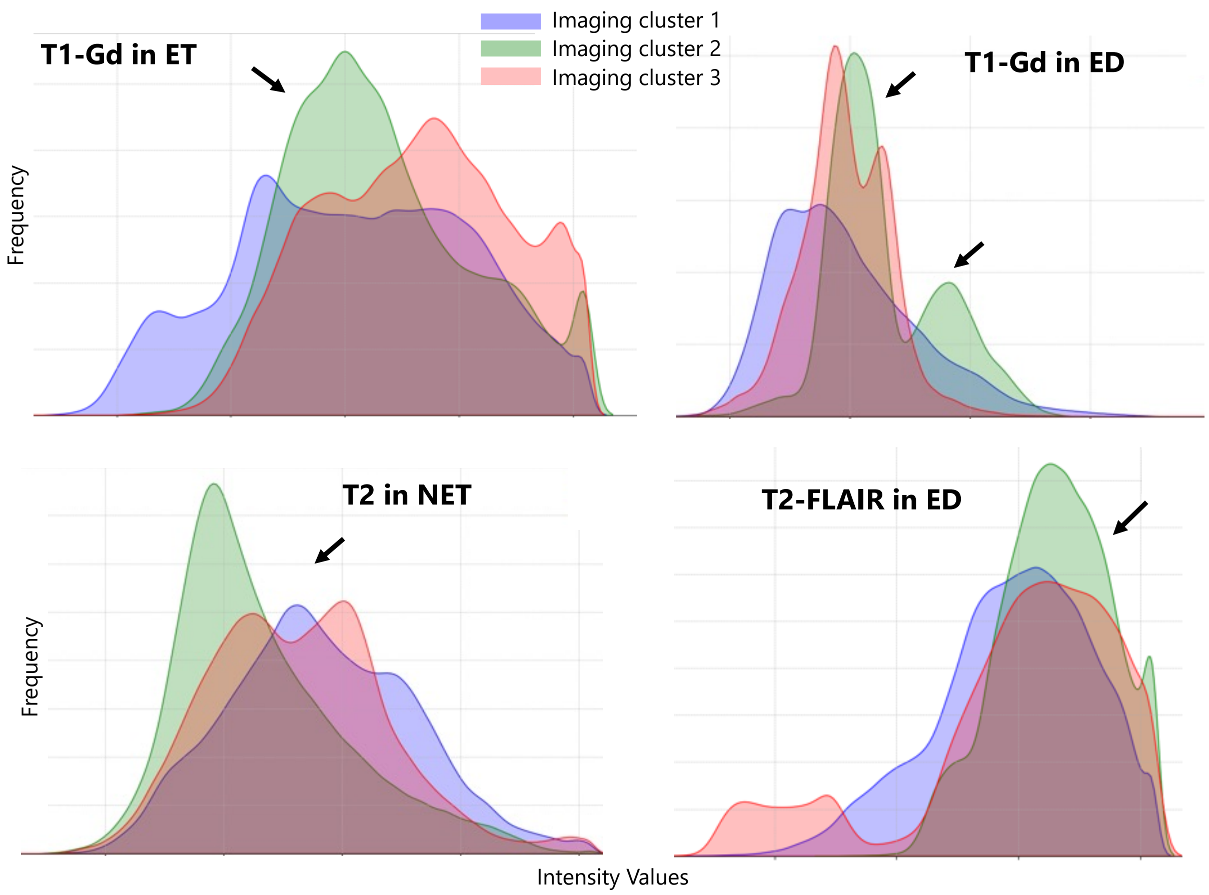


*Supplementary Figure 2. Histograms of image intensities within different tumorous subregions, i.e., enhancing tumor (ET), edema (ED), non-enhancing tumor (NET), for the three imaging clusters.*

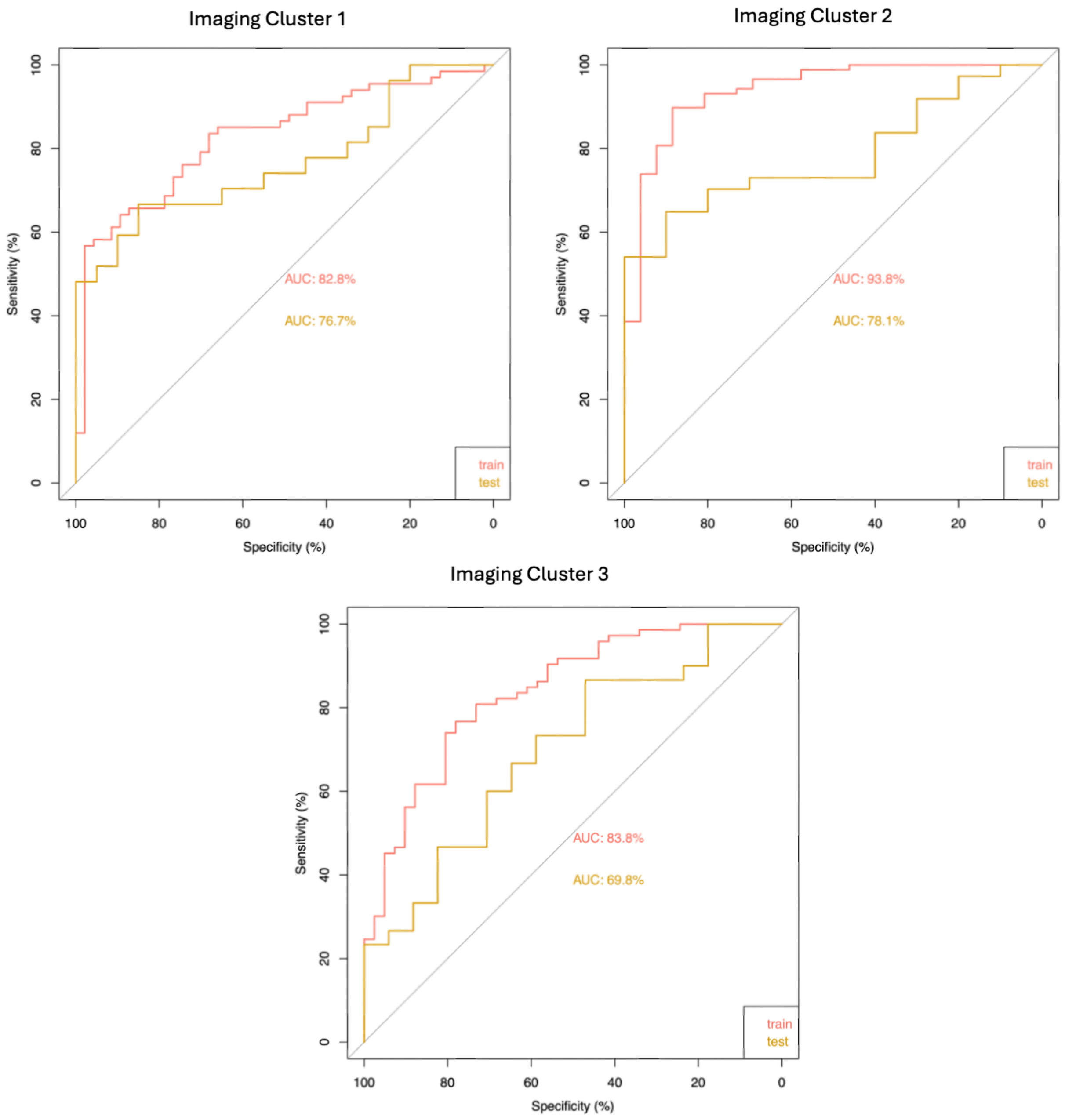


*Supplementary Figure 3. ROC curves illustrating the performances of elastic-net regression models applied to transcriptomic pathways to classify each imaging cluster from the rest. This analysis compares the relative association of differentially expressed/co-expressed signaling pathways, molecular subtypes, and clinical variables with imaging clusters.*

*Supplementary Table 2. Summary of Poisson generalized linear model parameters comparing counts across imaging clusters and molecular subtypes.*

| **Characteristic** | **IRR**[1] | **95% CI**[1] | **p-value** |
|---|---|---|---|
| 2021_WHO_Classification | | | |
| Pediatric-type diffuse low-grade gliomas, NOS | — | — | |
| Diffuse low-grade glioma, MAPK pathway-altered | 0.64 | 0.49, 0.83 | <0.001 |
| Ganglioglioma | 0.07 | 0.00, 0.32 | 0.008 |
| Pilocytic astrocytoma | 3.00 | 2.60, 3.49 | <0.001 |
| Imaging_Cluster | | | |
| 1 | — | — | |
| 2 | 0.36 | 0.23, 0.53 | <0.001 |
| 3 | 0.86 | 0.69, 1.06 | 0.2 |
| 2021_WHO_Classification * Imaging_Cluster | | | |
| Diffuse low-grade glioma, MAPK pathway-altered * 2 | 4.67 | 2.91, 7.75 | <0.001 |
| Ganglioglioma * 2 | 11.2 | 2.15, 207 | 0.022 |
| Pilocytic astrocytoma * 2 | 0.60 | 0.38, 0.98 | 0.034 |
| Diffuse low-grade glioma, MAPK pathway-altered * 3 | 2.85 | 2.08, 3.94 | <0.001 |
| Ganglioglioma * 3 | 3.50 | 0.65, 64.9 | 0.2 |
| Pilocytic astrocytoma * 3 | 0.50 | 0.39, 0.64 | <0.001 |

[1] IRR = Incidence Rate Ratio, CI = Confidence Interval

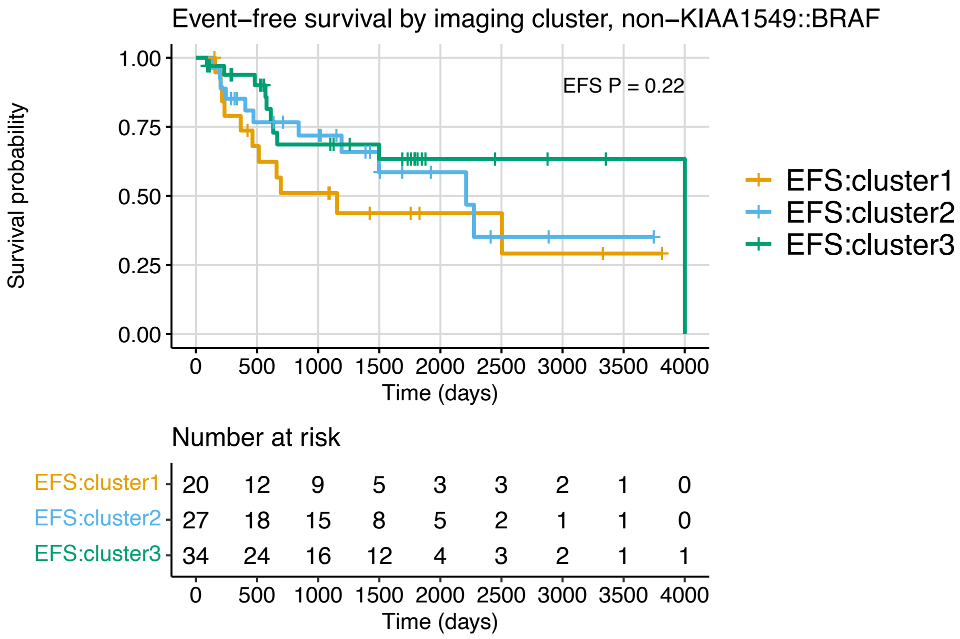


*Supplementary Figure 4. Kaplan Meier plots of event-free survival by imaging cluster in non-KIAA1549::BRAF fusion tumors.*

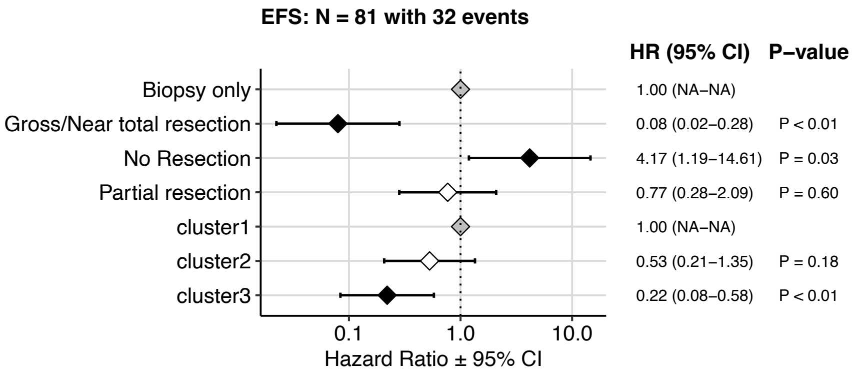


*Supplementary Figure 5. Cox regression model forest plots of event-free survival by imaging cluster in non-KIAA1549::BRAF fusion tumors including covariates for extent of tumor resection and imaging cluster.*

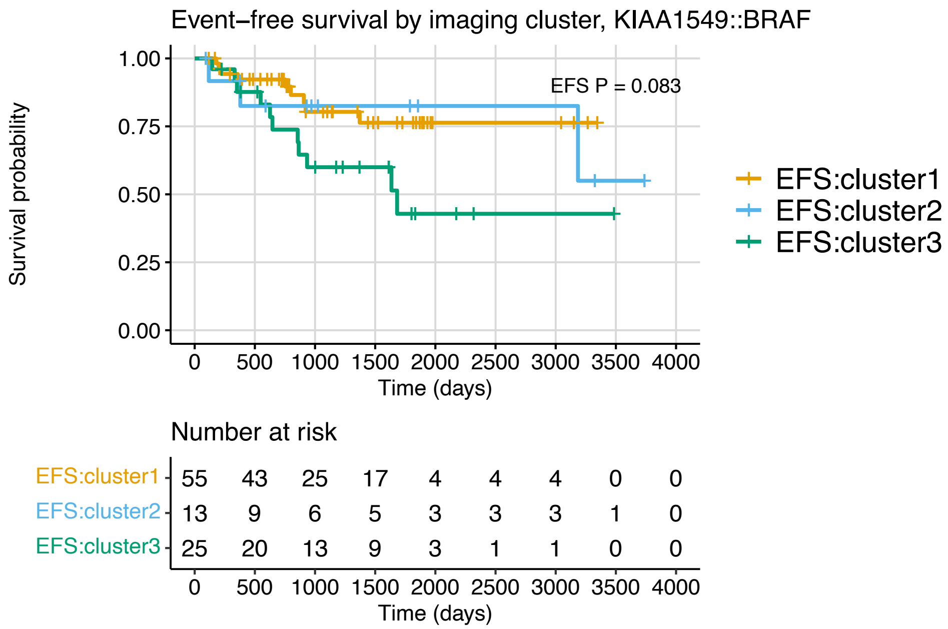


*Supplementary Figure 6. Kaplan Meier plots of event-free survival by imaging cluster in KIAA1549::BRAF fusion tumors.*

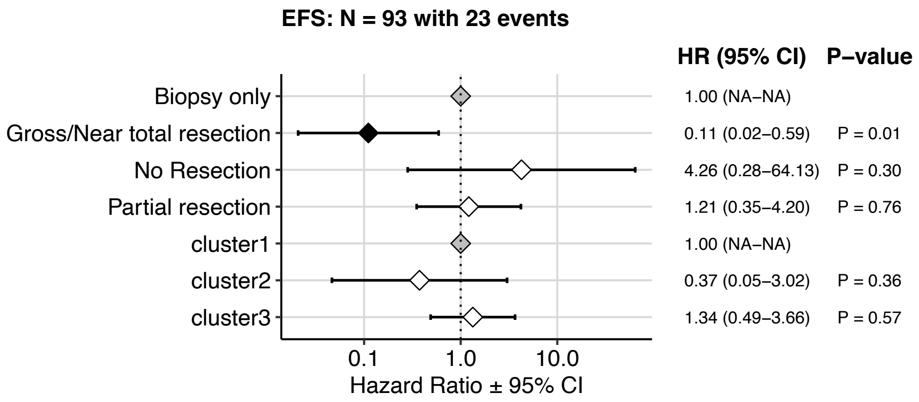


*Supplementary Figure 7. Cox regression model forest plots of event-free survival by imaging cluster in KIAA1549::BRAF fusion tumors including covariates for extent of tumor resection and imaging cluster.*

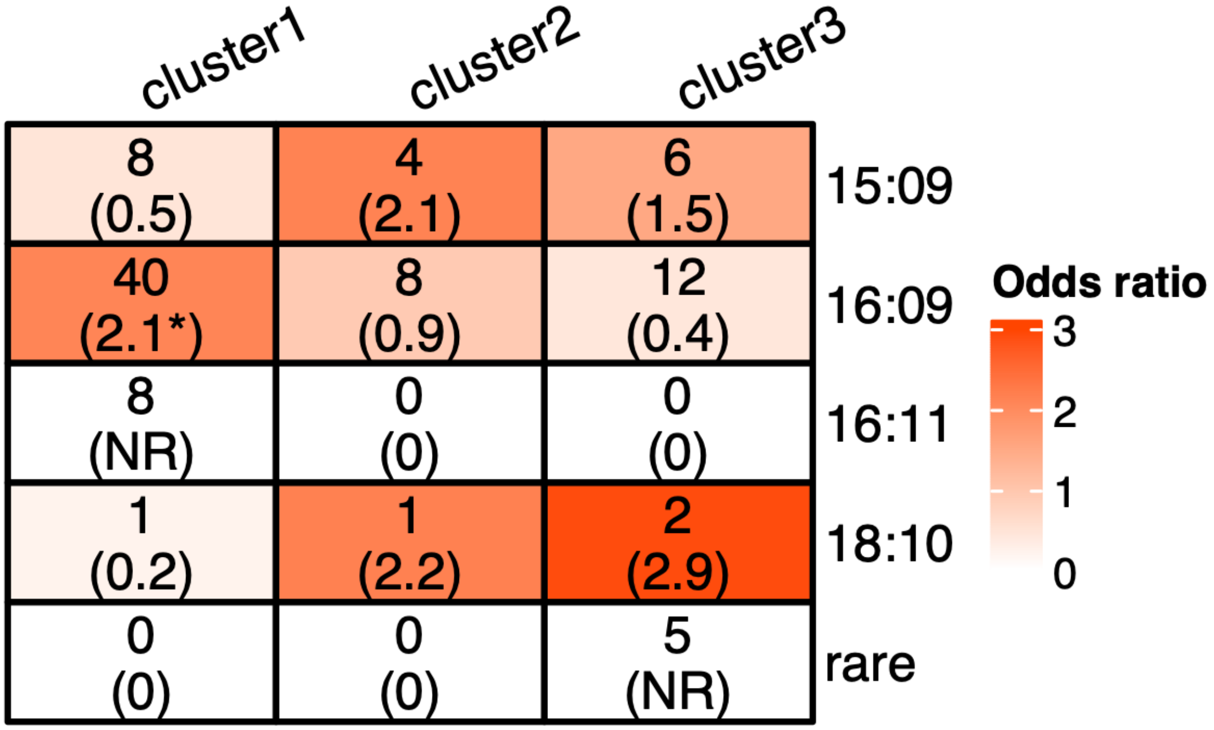


*Supplementary Figure 8. Distribution of KIAA1549::BRAF fusion tumor breakpoint groups among imaging clusters, and corresponding enrichment odds ratios in parentheses. *p<0.05. NR = Not reportable.*

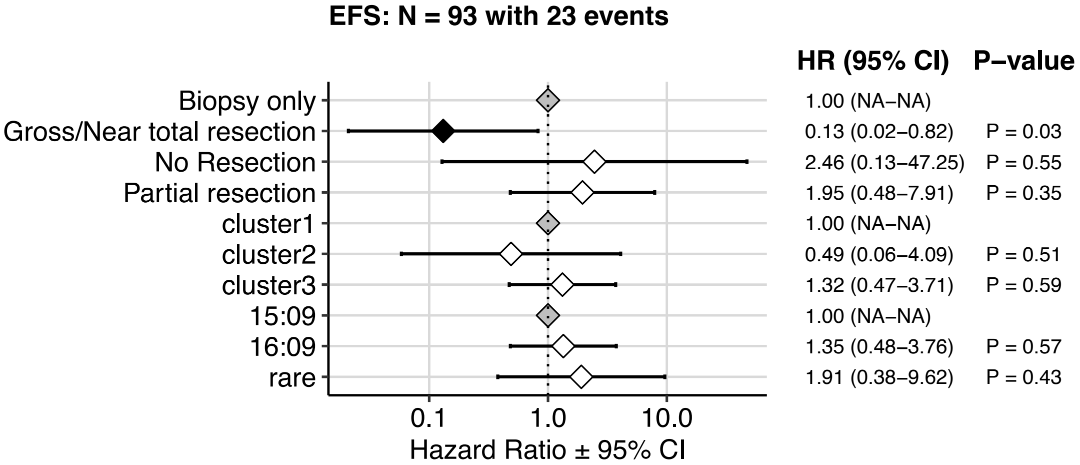


*Supplementary Figure 9. Cox regression model forest plot of EFS among patients with KIAA1549::BRAF fusion tumors, including covariates for extent of tumor resection, imaging cluster, and breakpoint group.*

*Gray points indicate reference levels for each covariate, and black points indicate terms with statistically significant hazard ratios relative to reference levels.*
